# Identification of biological correlates associated with respiratory failure in COVID-19

**DOI:** 10.1101/2020.09.29.20204289

**Authors:** Jung Hun Oh, Allen Tannenbaum, Joseph O Deasy

## Abstract

**Background:** Coronavirus disease 2019 (COVID-19) is a global public health concern. Recently, a genome-wide association study (GWAS) was performed with participants recruited from Italy and Spain by an international consortium group.

**Methods:** Summary GWAS statistics for 1610 patients with COVID-19 respiratory failure and 2205 controls were downloaded. In the current study, we analyzed the summary statistics with the information of loci and p-values for 8,582,968 single-nucleotide polymorphisms (SNPs), using gene ontology analysis to determine the top biological processes implicated in respiratory failure in COVID-19 patients.

**Results:** We considered the top 708 SNPs, using a p-value cutoff of 5×10^−5^, which were mapped to the nearest genes, leading to 144 unique genes. The list of genes was input into a curated database to conduct gene ontology and protein-protein interaction (PPI) analyses. The top ranked biological processes were wound healing, epithelial structure maintenance, muscle system processes, and cardiac-relevant biological processes with a false discovery rate < 0.05. In the PPI analysis, the largest connected network consisted of 8 genes. Through literature search, 7 out of the 8 genes were found to be implicated in both pulmonary and cardiac diseases.

**Conclusion:** Gene ontology and protein-protein interaction analyses identified cardio-pulmonary processes that may partially explain the risk of respiratory failure in COVID-19 patients.

## Background

Coronavirus disease 2019 (COVID-19) caused by a novel coronavirus (severe acute respiratory syndrome coronavirus 2, SARS-CoV-2) has resulted in a global pandemic with a rapidly developing global health and economic crisis [1]. Most people with COVID-19 are asymptomatic or experience only mild symptoms [2]. However, about 5% of patients infected with the coronavirus develop acute lung injury and acute respiratory distress syndrome, possibly leading to lethal lung damage and even death [3].

The most common reported comorbidities associated with poor outcomes in COVID-19 include hypertension, diabetes, cardiovascular disease, and chronic respiratory infections [4, 5]. However, the underlying molecular mechanisms in severe COVID-19 and their interplay with such comorbidities or clinical factors are poorly understood [6].

To identify putative biomarkers that can help better understand the molecular basis of COVID-19, Blanco-Melo *et al*. investigated the host transcriptional response to SARS-CoV-2 and other respiratory infections through *in vitro, ex vivo*, and *in vivo* experiments [1]. Bioinformatical approaches including gene ontology and protein-protein interaction (PPI) analyses were performed to identify key biological correlates. To investigate key genetic variants associated with respiratory failure in COVID-19 patients, a genome-wide association study (GWAS) was carried out on participants recruited from Italy and Spain [7]. In the current study, we performed an in-depth biological characterization including gene ontology and PPI analyses on summary statistics that resulted from the GWAS analysis in order to identify key biological correlates relevant to respiratory failure in COVID-19 patients.

## Methods

The GWAS conducted by an international consortium group involved 1980 patients with severe acute respiratory failure induced by COVID-19 at seven hospitals in Italy and Spain [7]. After quality control, the final case-control cohort included 835 patients and 1255 control participants from Italy and 775 patients and 950 control participants from Spain. After genotyping and imputation on genome build GRCh38, univariate analysis was performed for 8,582,968 single-nucleotide polymorphisms (SNPs). The resulting summary statistics including individual SNP positions and p-values were submitted to the European Bioinformatics Institute (www.ebi.ac.uk/gwas; accession numbers, GCST90000255 and GCST90000256) and are available from www.c19-genetics.eu. The GCST90000255 was a main analysis in which all the association statistics were corrected for the top 10 principal components (PCs), whereas in the additional analysis of GCST90000256, association statistics were corrected for the top 10 PCs, age, and sex. In the current study, we downloaded the summary statistics of GCST90000255 for further biological analysis.

To further enrich gene ontology terms with more plausible SNPs likely relevant to acute respiratory failure in COVID-19, we employed a relaxed p-value of 5×10^−5^ as a filtering threshold on the summary statistics. The SNPs with p-values < 5×10^−5^ were mapped to nearest genes using a 50kb window on both upstream and downstream sides of each gene. The resulting list of genes was together fed into MetaCore software (Thompson Reuters, New York, NY) for gene ontology analysis. Further PPI analysis was performed to explore the largest connected network, assuming that interacting proteins in a biological network may have the same or similar molecular functions [8-10].

## Results

Figure 1 shows quantile-quantile (QQ) and Manhattan plots for the GCST90000255. Strong deviation in the QQ plot indicates the possible association of SNPs with respiratory failure in COVID-19 patients. As shown in the Manhattan plot, there were two regions (loci 3p21.31 and 9q34.2) that reached a genome-wide significance level of 5×10^−8^.

**Figure 1.**
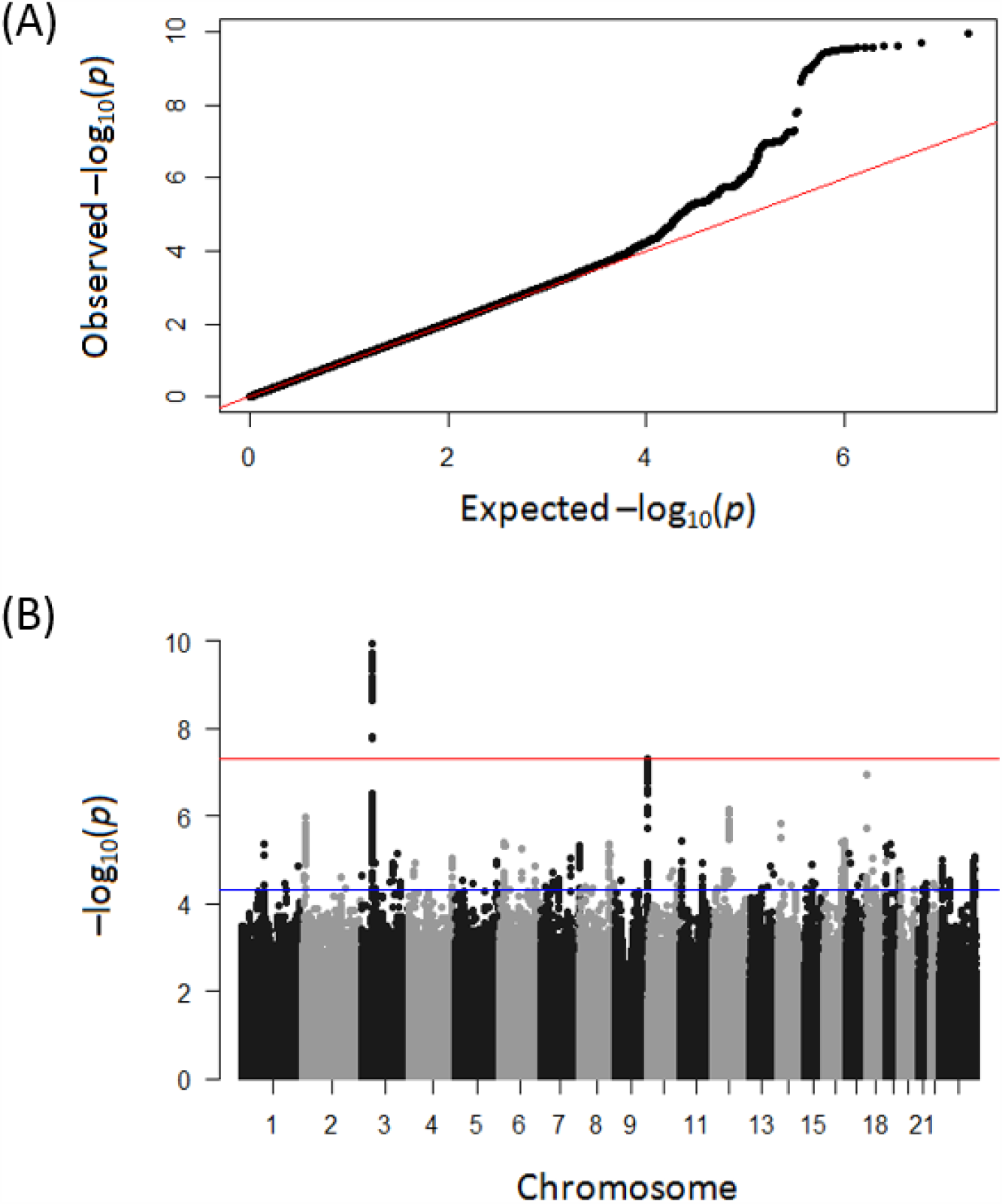
(A) quantile-quantile plot for the GCST90000255. The straight red line indicates the distribution under the null hypothesis that there is no association between any of the SNPs and respiratory failure in COVID-19 patients. (B) Manhattan plot. The red and blue lines indicate p-values of 5×10^−8^ and 5×10^−5^, respectively.

## Gene ontology analysis

In our analysis, with a p-value threshold of 5×10^−5^ applied to the summary statistics, 708 SNPs remained and a corresponding set of 144 unique genes in autosomes was found. The list of genes was fed into a MetaCore database. Table 1 shows the top 10 biological processes and corresponding genes that appear to be relevant to respiratory failure in COVID-19 patients, all with a false discovery rate (FDR) < 0.05. Wound healing, epithelial structure maintenance, muscle system process, and cardiac-relevant biological processes were top-ranked.

**Table 1.** The top 10 significant biological processes likely associated with respiratory failure in COVID-19 patients. The genes for each biological process belong to the list of 144 genes. FDR: false discovery rate.

| Ranking | Gene Ontology | FDR | Genes |
| --- | --- | --- | --- |
| 1 | Wound healing | 1.962E-2 | CCR9, CXCR6, TFF1, TFF3, MYH1, MYH4, ITGB3, SMAD3, GATA4, EPPK1, ADAMTS13, PRTN3, UBASH3A, MYH2, DMBT1, PLEC |
| 2 | Epithelial structure maintenance | 1.962E-2 | TFF1, TFF2, TFF3, LDB2 |
| 3 | Cardiac ventricle development | 1.962E-2 | CCR9, CXCR6, MYH1, MYH4, ID2, SLIT3, GATA4, SUFU, PTBP1 |
| 4 | Ventricular septum development | 1.962E-2 | CCR9, CXCR6, ID2, SLIT3, GATA4, SUFU |
| 5 | Cardiac septum development | 1.962E-2 | CCR9, CXCR6, MYH1, MYH4, ID2, SLIT3, GATA4, SUFU |
| 6 | Transdifferentiation | 1.962E-2 | SMAD3, GATA4 |
| 7 | Muscle system process | 2.106E-2 | CCR9, CXCR6, TMOD2, MYH1, MYH4, TMOD3, GATA4, MYH2, CHRNA1 |
| 8 | Cellular component maintenance | 2.518E-2 | CCR9, CXCR6, DLGAP1, PRTN3, TANC1, ERC2 |
| 9 | Embryonic foregut morphogenesis | 2.624E-2 | SMAD3, GATA4 |
| 10 | Response to virus | 2.783E-2 | CCR9, CXCR6, TRIM22, GATA4, TRIM5, TRIM34, DDX1, IL12A, DMBT1, AZU1, TRIM6 |

## Protein-protein interaction network analysis

The largest connected PPI network in the list of 144 genes is shown in Figure 2. The PPI network consisted of 8 genes: *GATA4, ID2, MAFA, NOX4, PTBP1, SMAD3, TUBB1*, and *WWOX*. We conducted a literature search in PubMed to investigate the potential associations between those 8 genes/proteins and pulmonary or cardiac diseases. Table 2 lists an overview of reported studies in terms of the associations. Interestingly, except for *MAFA* that is involved in insulin secretion, all 7 genes were found to be implicated in both pulmonary and cardiac diseases.

**Table 2.** An overview of reported studies in terms of the associations between genes/proteins and pulmonary or cardiac symptoms. The MAFA gene is involved in insulin secretion.

|  | Pulmonary symptoms | Cardiac symptoms | Insulin |
| --- | --- | --- | --- |
| GATA4 | Ackerman [11] | Garg [12]<br>Sarkozy [13] |  |
| ID2 | Arwood [17] | Jongbloed [18] |  |
| NOX4 | Guo [19] | Zhao [20] |  |
| SMAD3 | Gauldie [21] | Huang [22] |  |
| TUBB1 | Huang [23] | Freson [24] |  |
| PTBP1 | Caruso [25] | Fochi [26] |  |
| WWOX | Teixeira Gomes [27] | Tanna [28] |  |
| MAFA |  |  | Wang [29]<br>Zhang [30] |

**Figure 2.**
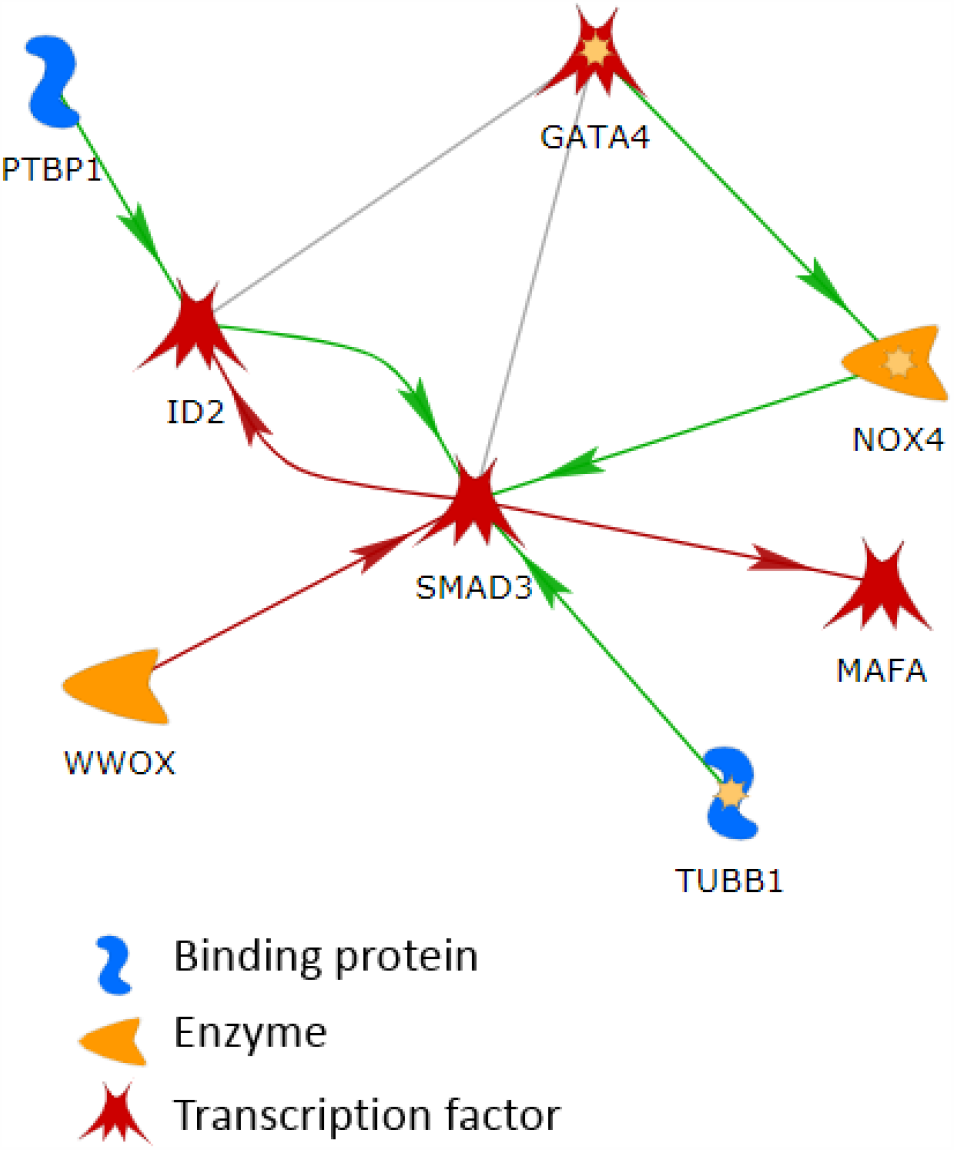
The largest connected network in the list of 144 genes. The line colors indicate the activation (green), inhibition (red), and unspecified (grey) effects.

## Discussion

Summary statistics from a GWAS for respiratory failure in COVID-19 patients was analyzed employing techniques from bioinformatics. To enrich biological discovery, a relaxed p-value of 5×10^−5^ was adopted, which likely enabled the inclusion of much potential genomic information in the analysis and the identification of plausible biological correlates associated with pulmonary or cardiac symptoms. A list of SNPs filtered by the relaxed p-value threshold was mapped to genes. The resulting 144 genes were fed into a MetaCore database for gene ontology and PPI analyses.

Gene ontology analysis identified wound healing, cardiac-related biological process, and muscle system process as key correlates. For PPI analysis, we attempted to find the largest connected network in the list of 144 genes, assuming that interacting proteins in a biological network tend to have the same or similar molecular functions. As a result, a network consisting of 8 genes was identified including *GATA4, ID2, MAFA, NOX4, PTBP1, SMAD3, TUBB1*, and *WWOX*. A literature search was conducted through PubMed to investigate whether there were previously reported results in terms of biological associations between these genes/proteins and respiratory or cardiac symptoms. Interestingly, we found that most of these genes are relevant to both respiratory and cardiac diseases. In what follows, we describe the role of these biomarkers in biology.

A study reported that *GATA4* plays a critical role as a transcription factor in normal pulmonary development [11]. *GATA4* also has been found to be a human candidate gene relevant to congenital heart disease [12, 13]. Several studies showed that *GATA4* is a key protein responsible for the development of the lung, heart, and diaphragm in mice [14-16].

Arwood *et al*. described a mechanism of pulmonary hypertension in heart failure with preserved ejection fraction (HFpEF) using transcriptome-wide RNA sequencing [17]. When comparing transcriptomics between patients with combined post- and pre-capillary pulmonary hypertension and those without pulmonary hypertension, six differentially expressed genes were identified. In a further replication test on an independent cohort, only *ID2* was validated and in an animal study, *ID2* expression was significantly upregulated in mice with HFpEF and pulmonary hypertension compared to control mice. Another study showed a functional role of *ID2* as one of the culprit genes in both the arterial and the venous poles of the heart [18].

An increased expression of *NOX4* and *TGF-β* was found to be correlated with the increased volume of airway smooth muscle mass and epithelial cells in small airways in the lung with chronic obstructive pulmonary disease (COPD) [19]. Another study reported that the upregulation of *NOX4* in heart induced cardiac remodeling, suggesting its potential to reduce the severity of established heart failure [20].

Gauldie *et al*. demonstrated the interactions between inflammation, *TGF-β* activation, and *SMAD3* signaling and pulmonary fibrosis and emphysema [21]. Huang *et al*. found that *SMAD3* is a key mediator in chronic cardiovascular disease, and plays a critical role in hypertensive cardiac remodeling [22].

At 4 and 24 hours after respiratory syncytial virus infection, gene expression profiles in human bronchial epithelial cells were analyzed [23]. Among the six genes that were associated with respiratory disease and were significantly altered at both 4 and 24 hours post-infection, *TUBB1* was the only gene observed to be downregulated at both time points. Freson *et al*. showed that the *TUBB1* Q43P functional variant may be a protective genetic factor against cardiovascular disease [24].

Caruso *et al*. observed the downregulation of *miR-124* in patients with pulmonary arterial hypertension and its central role in contributing to abnormal cell proliferation via *PTBP1* and *PKM2* [25]. Recently, Fochi *et al*. showed the emerging role of *RBM20* and *PTBP1* as key splicing factors in heart development and cardiovascular disease [26].

A study reported that loss of *WWOX* promoted cell proliferation in pulmonary artery smooth muscle cells and contributed to pulmonary vascular remodeling in pulmonary arterial hypertension [27]. Another study reported the vital implications of *WWOX* in atherosclerosis and cardiovascular diseases [28].

*MAFA* has not been found to be directly related to pulmonary or cardiac symptoms in the literature review. However, *MAFA* has been shown to be a key regulator that controls genes implicated in insulin secretion [29, 30]. A recent study indicated that a number of patients with COVID-19, who were comorbid with diabetes and several diabetes-related traits, were associated with increased *ACE2* expression [31]. This suggests that *ACE2* appears to be a potentially key molecular link between insulin resistance and COVID-19 severity [32].

This combined evidence indicated that lung disease is likely to be associated with cardiovascular risk. Further research should be warranted to identify the common biological processes between lung and heart diseases and the interplay between them.

We further assessed various filtering thresholds. With a stricter p-value of 1×10^−5^, 390 SNPs and corresponding 27 unique genes in autosomes remained. Gene ontology analysis with the relatively small number of genes resulted in immunity-related biological processes as the top important covariates. The top two biological processes were chemokine-mediated signaling pathway (FDR=2.906E-3) and CD8-positive, gamma-delta intraepithelial T cell differentiation (FDR=2.906E-3). With a more relaxed p-value of 1×10^−4^, 1112 SNPs and corresponding 243 unique genes in autosomes remained. Gene ontology analysis with those genes resulted in finding biological processes that are irrelevant to respiratory failure, which is likely due to false positives added in the analysis. This implies that the selection of an optimal threshold is critical to identify real biological correlates. Information informed by machine learning-based predictive modeling on GWAS data, which we employed in other studies [8, 9], can help resolve the issue.

## Conclusions

We analyzed summary statistics from a GWAS where individual SNPs were tested for associations with respiratory failure in COVID-19 patients. Bioinformatic approaches with SNPs filtered using a relaxed p-value enabled the identification of plausible biological correlates that are likely to be relevant to pulmonary or cardiac symptoms. When genotyping data become available, a more in-depth analysis using machine learning and bioinformatics techniques should provide greater insights into the underlying mechanisms of respiratory failure in COVID-19 patients.

## Data Availability

All the data analyzed in this study are available at http://www.c19-genetics.eu.

## Acknowledgements

This research was funded in part through the NIH/NCI Cancer Center Support Grant P30 CA008748 and R21 CA234752.

## Competing interests

The authors declare no conflicts of interest.

## Notes

### Competing Interest Statement

The authors have declared no competing interest.

## Reference

Blanco-Melo D, Nilsson-Payant BE, Liu WC, Uhl S, Hoagland D, Møller R, Jordan TX, Oishi K, Panis M, Sachs D et al: Imbalanced Host Response to SARS-CoV-2 Drives Development of COVID-19. Cell 2020, 181(5):1036-1045.e1039.

Yaqinuddin A, Kvietys P, Kashir J: COVID-19: Role of neutrophil extracellular traps in acute lung injury. Respir Investig 2020.

Baksh M, Ravat V, Zaidi A, Patel RS: A Systematic Review of Cases of Acute Respiratory Distress Syndrome in the Coronavirus Disease 2019 Pandemic. Cureus 2020, 12(5):e8188.

Di Carlo DT, Montemurro N, Petrella G, Siciliano G, Ceravolo R, Perrini P: Exploring the clinical association between neurological symptoms and COVID-19 pandemic outbreak: a systematic review of current literature. J Neurol 2020.

Malik YS, Kumar N, Sircar S, Kaushik R, Bhat S, Dhama K, Gupta P, Goyal K, Singh MP, Ghoshal U et al: Coronavirus Disease Pandemic (COVID-19): Challenges and a Global Perspective. Pathogens 2020, 9(7).

Kalfaoglu B, Almeida-Santos J, Adele Tye C, Satou Y, Ono M: T-cell hyperactivation and paralysis in severe COVID-19 infection revealed by single-cell analysis. bioRxiv 2020.

Ellinghaus D, Degenhardt F, Bujanda L, Buti M, Albillos A, Invernizzi P, Fernández J, Prati D, Baselli G, Asselta R et al: Genomewide Association Study of Severe Covid-19 with Respiratory Failure. N Engl J Med 2020.

Oh JH, Kerns S, Ostrer H, Powell SN, Rosenstein B, Deasy JO: Computational methods using genome-wide association studies to predict radiotherapy complications and to identify correlative molecular processes. Sci Rep 2017, 7:43381.

Lee S, Kerns S, Ostrer H, Rosenstein B, Deasy JO, Oh JH: Machine Learning on a Genome-wide Association Study to Predict Late Genitourinary Toxicity After Prostate Radiation Therapy. Int J Radiat Oncol Biol Phys 2018, 101(1):128–135.

Lee S, Liang X, Woods M, Reiner AS, Concannon P, Bernstein L, Lynch CF, Boice JD, Deasy JO, Bernstein JL et al: Machine learning on genome-wide association studies to predict the risk of radiation-associated contralateral breast cancer in the WECARE Study. PLoS One 2020, 15(2):e0226157.

Ackerman KG, Wang J, Luo L, Fujiwara Y, Orkin SH, Beier DR: Gata4 is necessary for normal pulmonary lobar development. Am J Respir Cell Mol Biol 2007, 36(4):391–397.

Garg V, Kathiriya IS, Barnes R, Schluterman MK, King IN, Butler CA, Rothrock CR, Eapen RS, Hirayama-Yamada K, Joo K et al: GATA4 mutations cause human congenital heart defects and reveal an interaction with TBX5. Nature 2003, 424(6947):443–447.

Sarkozy A, Conti E, Neri C, D’Agostino R, Digilio MC, Esposito G, Toscano A, Marino B, Pizzuti A, Dallapiccola B: Spectrum of atrial septal defects associated with mutations of NKX2.5 and GATA4 transcription factors. J Med Genet 2005, 42(2):e16.

Jay PY, Bielinska M, Erlich JM, Mannisto S, Pu WT, Heikinheimo M, Wilson DB: Impaired mesenchymal cell function in Gata4 mutant mice leads to diaphragmatic hernias and primary lung defects. Dev Biol 2007, 301(2):602–614.

Olson EN: Gene regulatory networks in the evolution and development of the heart. Science 2006, 313(5795):1922–1927.

Kimura Y, Suzuki T, Kaneko C, Darnel AD, Moriya T, Suzuki S, Handa M, Ebina M, Nukiwa T, Sasano H: Retinoid receptors in the developing human lung. Clin Sci (Lond) 2002, 103(6):613– 621.

Arwood MJ, Vahabi N, Lteif C, Sharma RK, Machado RF, Duarte JD: Transcriptome-wide analysis associates ID2 expression with combined pre- and post-capillary pulmonary hypertension. Sci Rep 2019, 9(1):19572.

Jongbloed MR, Vicente-Steijn R, Douglas YL, Wisse LJ, Mori K, Yokota Y, Bartelings MM, Schalij MJ, Mahtab EA, Poelmann RE et al: Expression of Id2 in the second heart field and cardiac defects in Id2 knock-out mice. Dev Dyn 2011, 240(11):2561–2577.

Guo X, Fan Y, Cui J, Hao B, Zhu L, Sun X, He J, Yang J, Dong J, Wang Y et al: NOX4 expression and distal arteriolar remodeling correlate with pulmonary hypertension in COPD. BMC Pulm Med 2018, 18(1):111.

Zhao QD, Viswanadhapalli S, Williams P, Shi Q, Tan C, Yi X, Bhandari B, Abboud HE: NADPH oxidase 4 induces cardiac fibrosis and hypertrophy through activating Akt/mTOR and NFκB signaling pathways. Circulation 2015, 131(7):643–655.

Gauldie J, Kolb M, Ask K, Martin G, Bonniaud P, Warburton D: Smad3 signaling involved in pulmonary fibrosis and emphysema. Proc Am Thorac Soc 2006, 3(8):696–702.

Huang XR, Chung AC, Yang F, Yue W, Deng C, Lau CP, Tse HF, Lan HY: Smad3 mediates cardiac inflammation and fibrosis in angiotensin II-induced hypertensive cardiac remodeling. Hypertension 2010, 55(5):1165–1171.

Huang YC, Li Z, Hyseni X, Schmitt M, Devlin RB, Karoly ED, Soukup JM: Identification of gene biomarkers for respiratory syncytial virus infection in a bronchial epithelial cell line. Genomic Med 2008, 2(3-4):113–125.

Freson K, De Vos R, Wittevrongel C, Thys C, Defoor J, Vanhees L, Vermylen J, Peerlinck K, Van Geet C: The TUBB1 Q43P functional polymorphism reduces the risk of cardiovascular disease in men by modulating platelet function and structure. Blood 2005, 106(7):2356–2362.

Caruso P, Dunmore BJ, Schlosser K, Schoors S, Dos Santos C, Perez-Iratxeta C, Lavoie JR, Zhang H, Long L, Flockton AR et al: Identification of MicroRNA-124 as a Major Regulator of Enhanced Endothelial Cell Glycolysis in Pulmonary Arterial Hypertension via PTBP1 (Polypyrimidine Tract Binding Protein) and Pyruvate Kinase M2. Circulation 2017, 136(25):2451–2467.

Fochi S, Lorenzi P, Galasso M, Stefani C, Trabetti E, Zipeto D, Romanelli MG: The Emerging Role of the RBM20 and PTBP1 Ribonucleoproteins in Heart Development and Cardiovascular Diseases. Genes (Basel) 2020, 11(4).

Teixeira Gomes M, Chen J, Haider S, Bai Y, Singla S, Machado RF: Smooth Muscle Cell Loss of the Tumor Suppressor WWOX Contributes to the Development of Pulmonary Hypertension. Am J Respir Crit Care Med 2020, 201:A7211.

Tanna M, Aqeilan RI: Modeling WWOX Loss of Function in vivo: What Have We Learned? Front Oncol 2018, 8:420.

Wang H, Brun T, Kataoka K, Sharma AJ, Wollheim CB: MAFA controls genes implicated in insulin biosynthesis and secretion. Diabetologia 2007, 50(2):348–358.

Zhang C, Moriguchi T, Kajihara M, Esaki R, Harada A, Shimohata H, Oishi H, Hamada M, Morito N, Hasegawa K et al: MafA is a key regulator of glucose-stimulated insulin secretion. Mol Cell Biol 2005, 25(12):4969–4976.

Rao S, Lau A, So HC: Exploring Diseases/Traits and Blood Proteins Causally Related to Expression of ACE2, the Putative Receptor of SARS-CoV-2: A Mendelian Randomization Analysis Highlights Tentative Relevance of Diabetes-Related Traits. Diabetes Care 2020, 43(7):1416–1426.

Finucane FM, Davenport C: Coronavirus and Obesity: Could Insulin Resistance Mediate the Severity of Covid-19 Infection? Front Public Health 2020, 8:184.

